# Training and Development Needs Assessment in a large NIHR Biomedical Research Centre: A Survey

**DOI:** 10.1101/2021.08.27.21261708

**Authors:** Karen Bell, Syed Ghulam Sarwar Shah, Lorna R. Henderson, Vasiliki Kiparoglou

## Abstract

**Objective:** To assess the training and development needs of researchers and support staff affiliated to the NIHR Oxford Biomedical Research Centre (BRC), one of the largest BRCs in England, and to find out about their past experiences of training.

**Design:** A cross-sectional online questionnaire survey.

**Setting and Participants:** A convenience sample of clinicians, nurses, midwives, allied health professionals, researchers and support staff (N=798) affiliated with the NIHR Oxford Biomedical Research Centre.

**Primary and secondary outcome measures:** The primary outcome measure was the type of training and the secondary outcome measures were the duration, location and timing of training.

**Results:** The response rate was 24%. Of 189 respondents, 114 were women (60%) and 75 men (40%). Respondents included research scientists (31%), medical doctors and dentists (17%), nurses and midwives (16%) and research managers and administrators (16%). Seventy-one percent respondents (n=134) reported attending at least one training activity in the last year and the most wanted training was leadership skills (25%), followed by research grant and fellowship writing (18%) and statistical analysis (16%). An ideal length of a training course was half a day (41%), whole day (25%) and 1-2 hours (22%). The most preferred time of the day for training was morning (60%) and afternoon (22%) and the favoured delivery style of training was an interactive workshop (52%), lecture/talk (25%), online (9%) and practical activities (9%). The main barriers to attending training courses were the lack of time (n-18%), work commitments (13%), and childcare responsibilities (6%).

**Conclusions:** Translational researchers and supporting affiliates want short, easily accessible, interactive training sessions, particularly leadership training skills and grant and fellowship writing. However, practical elements are important too e.g. in a convenient location during the working day. Work commitment is the biggest obstacle in doing training.

**Strengths and limitations of this study:**

- This survey was done to develop and revamp the NIHR Oxford BRC’s training programme that met the training and development needs of our researchers and research support staff.
- Leadership skills, research grant and fellowship writing, statistical analysis were the most wanted training.
- The lack of time, work commitments, and childcare responsibilities were the main barriers to attending training courses.
- Our findings have limited generalisability because the study is based on the responses of participants who are affiliated with only one NIHR BRC; hence, these findings could not be generalised to other NIHR BRCs.
- These findings might inform the training and development programmes in other NIHR BRCs in the country.

## INTRODUCTION

The National Institute for Health Research (NIHR) Biomedical Research Centres are part of the Government’s initiative to improve the translation of basic scientific developments into clinical benefits for patients and to reinforce the position of the UK as a global leader in healthcare related research.^1^ The NIHR Oxford Biomedical Research Centre (BRC) is a partnership that brings together the research expertise of the University of Oxford and the clinical skills of staff of Oxford University Hospitals NHS Foundation Trust with the aim of supporting translational research and innovation to improve healthcare for patients.^2^

The NIHR Oxford BRC ‘s overarching strategy focuses on building capacity with the explicit aim to attract, develop and retain the best research professionals.^1^ Firstly, by providing opportunities for talented healthcare research staff to undertake their own research through higher degrees, as well as via shorter research fellowships. Secondly, to facilitate the training and engagement in professional development of all of its affiliates including researchers and research support staff.

NIHR reviewed their training program in 2015^1^ and found that there was a need to develop innovative approaches to train the translational research workforce of the future, and to develop their career pathways as the clinical and translational environment is changing rapidly. Training is vital to maintain a skilled workforce as healthcare changes with technological advances and emerging diseases such as COVID-19.^3^ It is also a way for individuals to develop their careers, improving confidence and motivation and ultimately retention.^4^ In addition, training and development is essential for improving patient care^5^ as well as research and innovation.^6,7^

The NIHR Oxford BRC spends in the region of £300,000 a year on training and education for translational research staff. This is about 1.3% of its total annual budget of £23m. On average, about 70 researchers a year benefit from training support which includes providing training bursaries, fellowships, and bespoke courses including leadership, health economics and grant writing skills. The NIHR Oxford BRC works collaboratively with other organisations including the Clinical Research Network, the Oxford Health BRC and University of Oxford who also provide a range of free training opportunities for their students and staff.

Following reorganisation within the NIHR Oxford BRC a dedicated Training and Education Manager was appointed to a new role in 2019. In order to plan an effective training programme, we sought the views of researchers and support staff within and affiliated to the NIHR Oxford BRC about their training needs. This was imperative because according to the 2019 Researcher Development Concordat^8^, researchers must be equipped and supported to be adaptable and flexible in an increasingly diverse global research environment and employment market. This Principle recognises the importance of continuous professional and career development, particularly as researchers pursue a wide range of careers ^8^. Most clinical practitioners receive regular professional training such as good clinical practice and obtaining ethical approval, but not leadership training and research skills, which are associated with progression in rank, leadership position and research publication.^9^

The objective of this study was to assess the training and development needs of researchers and support staff affiliated to the NIHR Oxford BRC and to find out about their past experiences of training.

## METHODS

### Study Design

This was a cross sectional questionnaire survey study.

### Study setting

The NIHR Oxford BRC is based at the Oxford University Hospitals NHS Foundation Trust and run in partnership with the University of Oxford.^2^ Founded in 2007, it is one of five centres funded by the NIHR and has received over £260m since 2007 to support translational research. The NIHR Oxford BRC is divided into 20 research themes with over 500 research and research support staff paid for by the NIHR Oxford BRC.^2^ In addition to BRC staff, we also sent the questionnaire to people who are involved with translational research but not paid directly by the BRC, such as NHS research nurses.

### Study population

The study population included anyone involved in translational research and affiliated to the NIHR Oxford BRC. This included medical doctors, nurses, midwives, allied health professionals, clinical scientists, statisticians, software engineers, admin staff and clinical trial managers but was not limited to staff paid directly by the NIHR Oxford BRC.

### Development of the survey questionnaire

The survey questionnaire was developed in house and comprised 10 questions with a mix of multiple-choice questions and free text answers (**Appendix 1**. NIHR Oxford BRC Training and Development Questionnaire). These questions asked for participants’ gender and role, training type, time, duration, location and delivery style, training attended in the last year, most wanted training, and barriers to attending training. Participants were also given an open-ended choice to comment on the training received in the past. The questionnaire was intended to be quick and easy to complete while capturing the information needed to develop and revamp the NIHR Oxford BRC’s training programme that met the training and development needs of our researchers and research support staff. The questionnaire was developed using the JISC online survey software (JISC®) - an online survey tool designed for academic research, education and public sector organisations.^10^ The questionnaire was piloted with five members of the core administration team of the BRC.

### Administration of the survey

Using the JISC® online surveys^10^, the training survey was sent via personalised emails to 798 people associated/affiliated with the NIHR Oxford BRC in October 2019. They were given two months to respond, with two reminders. We received 189 responses (24%) by 31 December 2019. Responses were collated using the JISC software. With the JISC® online survey, data was secure and strict information security standards were followed (ISO27001)^11^ in compliance with the General Data Protection Regulations (GDPR).^12^

### Data analysis

Online data from JISC® was analysed and tables and graphs were produced for preliminary analysis. Data were also downloaded into SPSS^13^ and Microsoft Excel spreadsheet formats. Data were analysed for frequencies and descriptive statistics. We did not impute missing values, which were very low and we did not run any inferential statistical analyses. We did not conduct any sensitivity analysis.

### Patient and public involvement

Neither patients nor the public were involved in this study because it was a survey about the professional training and development needs of translational health researchers and support staff affiliated with our BRC.

### Reporting checklist

We report this study according to the Consensus-Based Checklist for Reporting of Survey Studies (CROSS)^14^.

## RESULTS

The response rate was 24% (189 responded of 798 invited). Of the 189 respondents, 114 were women (60%) and 75 were men (40%) and included research scientists, medical doctors and dentists, nurses and midwives, research managers and administrators and others (**Table 1**). The participants were from all 20 research themes plus the NIHR Oxford BRC management team (Table 1).

**Table 1.** Respondents’ role and training received and needed.

|  | Count | % |
| --- | --- | --- |
| **Participants' role (N=189)** |  |  |
| Research scientist | 59 | 31.2 |
| Medical doctor/dentist | 32 | 16.9 |
| Research nurse/midwife | 30 | 15.9 |
| Administrator/manager | 30 | 15.9 |
| Other (clinical scientists, pharmacists, statisticians) | 24 | 12.7 |
| Research Allied Health Professionals | 14 | 7.4 |
| **Training received in the last year\*** |  |  |
| Professional qualifications including masters' degrees | 23 | 12.2 |
| Good Clinical Practice (GCP) | 19 | 10.0 |
| Leadership | 14 | 7.4 |
| Bioinformatics and statistics | 12 | 6.3 |
| Clinical trials | 8 | 4.2 |
| Good Research Practice (GRP) | 8 | 4.2 |
| IT modules | 8 | 4.2 |
| Conferences and seminars | 7 | 3.7 |
| Ethics | 4 | 2.1 |
| General Data Protection Regulations (GDPR) | 4 | 2.1 |
| Grants (writing and applying) | 4 | 2.1 |
| Informed consent | 4 | 2.1 |
| Communications | 3 | 1.6 |
| Data management | 3 | 1.6 |
| Publications | 3 | 1.6 |
| Qualitative interviewing | 3 | 1.6 |
| Anti-bullying | 1 | 0.5 |
| Equality & diversity | 1 | 0.5 |
| Experimental design | 1 | 0.5 |
| Health Economics | 1 | 0.5 |
| Patient and Public Involvement (PPI) in Research | 1 | 0.5 |
| Project management | 1 | 0.5 |
| Teaching | 1 | 0.5 |
| None | 76 | 40.2 |
| **Training found most valuable (n=65)** |  |  |
| Leadership | 11 | 16.9 |
| Statistics | 8 | 12.3 |
| Good Clinical Practice (GCP) | 5 | 7.7 |
| Conferences | 5 | 7.7 |
| Information Technology (IT) | 5 | 7.7 |
| Lab/research skills | 5 | 7.7 |
| Clinical trials | 4 | 6.2 |
| Grants | 4 | 6.2 |
| Professional qualifications | 3 | 4.6 |
| Ethics | 3 | 4.6 |
| Publications | 3 | 4.6 |
| Networking | 2 | 3.1 |
| Epidemiology | 1 | 1.5 |
| Good Research Practice (GRP) | 1 | 1.5 |
| Clinical skills | 1 | 1.5 |
| Qualitative interviews | 1 | 1.5 |
| Informed Consent | 1 | 1.5 |
| Presentations | 1 | 1.5 |
| Teaching | 1 | 1.5 |
| **Training area most important to respondent's development and training needs (n=178)** |  |  |
| Leadership skills | 44 | 24.7 |
| Grant/fellowship writing | 32 | 18.0 |
| Statistical analysis | 28 | 15.7 |
| Clinical skills (PPI, GCP, informed consent) | 16 | 9.0 |
| Designing and conducting clinical trials | 16 | 9.0 |
| Other (Project management, programming and IT) | 15 | 8.4 |
| Research skills (GRP, lab skills) | 13 | 7.3 |
| Academic writing | 9 | 5.1 |
| Presentation skills | 5 | 2.8 |
| * Some respondents attended more than one training in the last year, so the total exceeds the number of total respondents (N=189, 100%) |  |  |

### Training received in the last year

Sixty percent of respondents (n=113) reported receiving at least one training course in the last year (**Table 1**). The most common training received included professional qualifications such as Master’s degree (n=23), followed by good clinical practice (n=19), leadership (n=14), and bioinformatics and statistics (n=12). However, about 40% respondents (n=76) reported not receiving any training in the last year.

### Most important training area for personal development and training

The respondents reported different areas of training for personal development and training and the most wanted training was leadership skills (25%), research grant and fellowship writing (18%) and statistical analysis (16%) (**Table 1**).

### Length and time of training

For respondents, the ideal length of a training course was half a day (41%), whole day (25%) and 1-2 hours (22%) (**Figure 1**). The most preferred time of the day for training was morning (60%) and afternoon (22%) (**Figure 1**).

**Figure 1.**
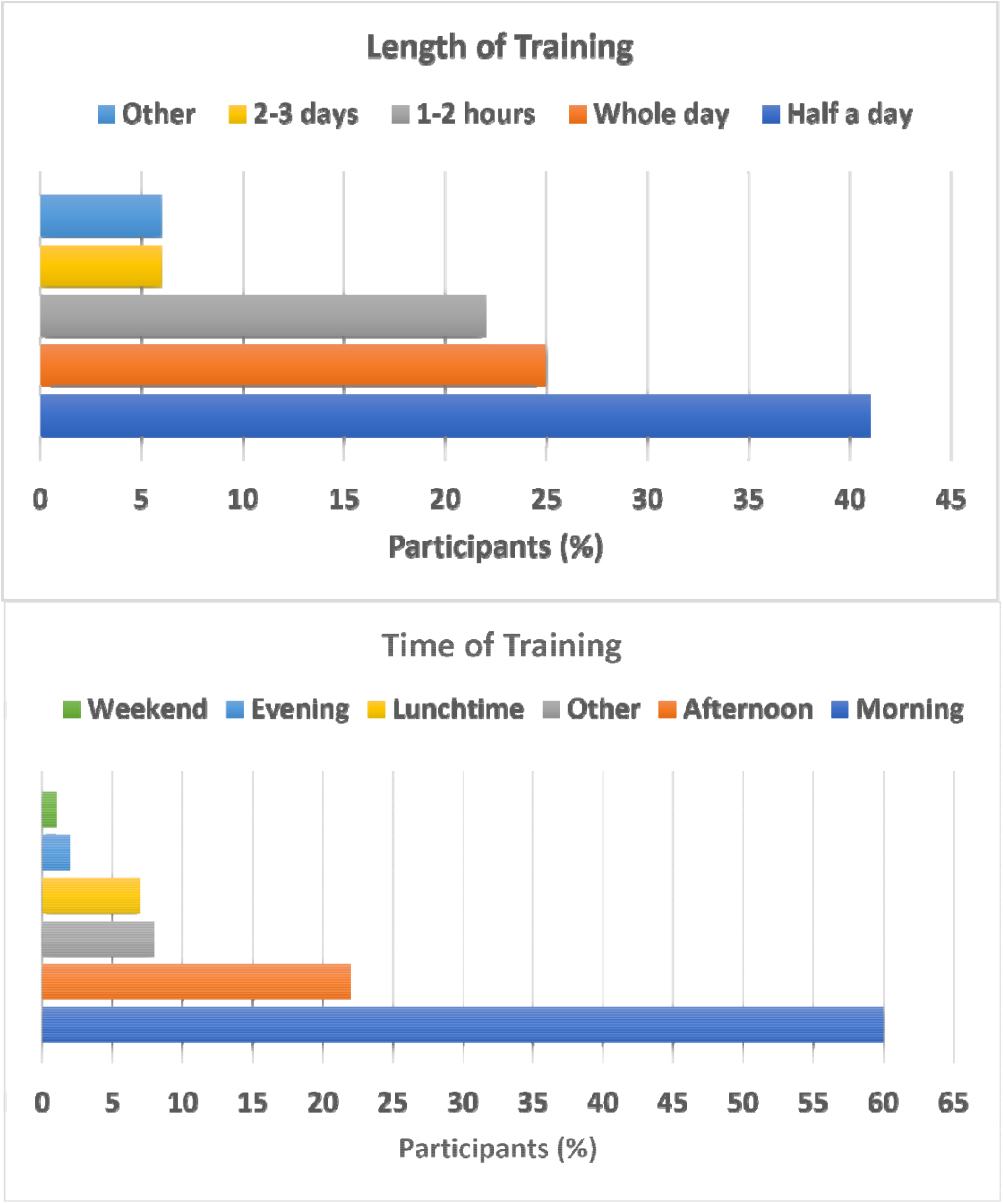
Ideal length and time of training.

### Training delivery

For the delivery of training, the participants suggested different delivery styles including interactive workshops (52%), lectures/talks (25%), online training (9%) and practical activities (9%) (**Figure 2**).

**Figure 2.**
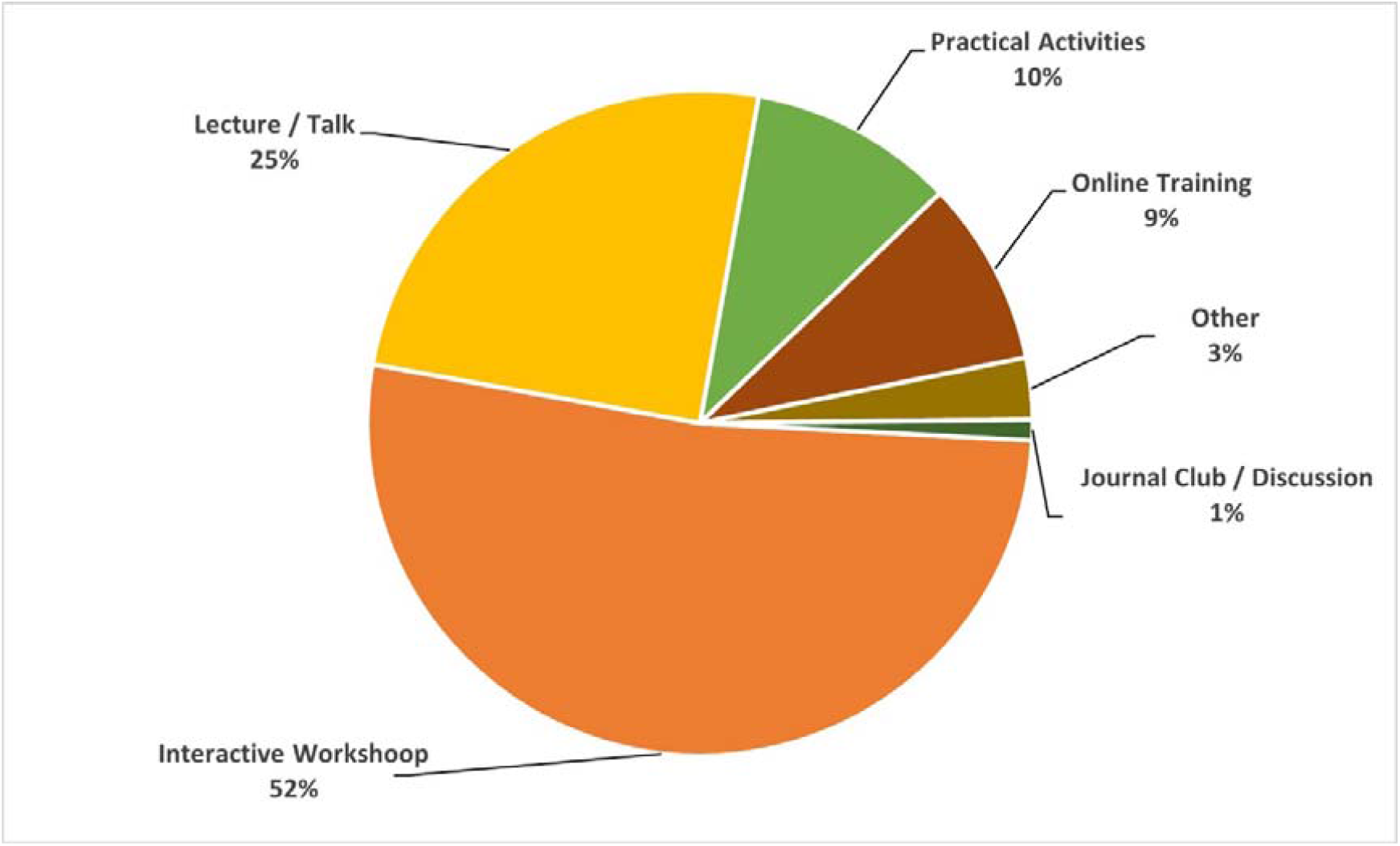
Training delivery styles suggested by participants.

### Barriers in attending Training

For 58% respondents (n=109) there were no barriers while the remaining participants (42%, n=80) reported a range of barriers that prevented them from attending training courses. The most common issues stopping from attending training courses were the lack of time (18%, n=34), work commitments (13%, n=25), and childcare responsibilities (6%, n=11). Other less common barriers to training were cost (2%, n=3), sufficient notice (1%, n= 2), permission (1%, n= 2), parking (n=1, 0.5%), location (n=1, 0.5%) and relevance (n=1, 0.5%).

### Open ended comments about training and development

Although a large number of respondents did not do any training (n=76, 40%), those who did training, found it very useful. **Box 1** provides selected comments showing the value and application of training in work, training tailored to the roles, research specific training, and training on clinical and epidemiological skills.

#### Box 1.

**Open ended comments by respondents**.

##### Training applicable at work

*The Biomedical Data science training program was an incredible course, I learned a lot and have been able to apply it to my own data*.*(Respondent # 34,Female, Research Scientist)*

##### Training tailored to the role

*The EMBO course: 30 hours of high quality leadership training tailored specifically to my role as a new PI. (Respondent # 35, Male, Research Scientist)*

*Leadership training SBS access to wide network of International leaders and techniques to apply to the BRC and make a difference*.*(Respondent # 43, Female, Manger)*

*Information Governance at HTA as these are key aspects of my role. (Respondent # 150, Female, research nurse/midwife)*

##### Research specific training

*Clinical Trial (training)…*.*allowed me to run my study more carefully [Respondent # 67, Male, medical doctor/ dentist]*

*Publication schools - excellent and engaging faculty, gained a lot of knowledge about publishing process and the university regulations. (Respondent # 47, Female, Medical doctor/ Dentist)*

*HRA approvals as it was short so able to go in work time and relevant to job. (Respondent # 129, Female, Research nurse/Midwife)*

*GCP update as relevant and practical and opportunity to meet other research staff. (Respondent # 134, Female, Research nurse/Midwife)*

##### Clinical and epidemiological skills training

*Epidemiological assessment of vaccines provided me with knowledge applicable to my current post. Tropical Nursing provided me with a wider knowledge of the diseases we are looking vaccines for. (Respondent # 78, Male, Research nurse / Midwife)*

*The vaccinology courses gave me more knowledge and understanding to work at a higher standard that was required, and the other training was helpful for career progressing*.*(Respondent # 185, Female, Administrator / manger)*

*Communications course - Extremely useful for having difficult conversations. (Respondent # 112, Female, medical doctor/dentist)*

## DISCUSSION

Nearly 800 people were personally sent the questionnaire of whom 54% were female and 46% male. The response rate was 24%, with six out of ten respondents being women (27% of cohort) and 40% men (20% of the cohort). The largest professional group to respond were research scientists at just over 30%.

Although, the majority of respondents reported receiving training in the past year, the usefulness of the training was mixed. However, training that was directly linked to professional and career development was well received because it has implications not only for developing competency, recruitment and retention but also improving healthcare delivery. ^4^ Training in leadership skills and research and grant writing were highlighted as possible future training opportunities because these skills are associated with progression in rank, leadership position and research publication. ^9^ In addition, training in leadership and management helps in increasing personal effectiveness and promoting a positive attitude to professional development. ^8^ Most notably, the need for leadership skills training was more in women compared to men respondents (7 women vs. 3 men). These findings indicate a gap in leadership in women in translational research settings. ^15,16^ Leadership is a marker of achievement in biomedical research organisations. ^17^ Therefore, gender equity in leadership is essential, ^16,17^ and the gender gap in leadership could be reduced by providing training in leadership skills.

Participants were also asked about the location, time and delivery of training courses. The most popular time of training delivery was mornings, training delivery style was interactive workshops and training location was near to where they work, which provides support to the recommendations made for the training of health and care workforce. ^18^ Our findings show that busy people, like clinicians and nurses, need to be able to access training easily, it has to be close by their workplace and to fit in with crowded work schedules. ^19^

A range of barriers to attending training were reported and top three barriers were the lack of time, work load and commitments and childcare responsibilities. ^20^ About half of nurses and medical doctors found time and work commitments a major barrier to training compared to a quarter of administrators, managers and AHPs and 15% of research scientists. Transportation and Parking, especially parking for people with disabilities, is a big issue in Oxford, so not having to travel is a bonus. These findings suggest that professional training must be inclusive and should take in to account the participants’ access, location, timing, physical limitations and family commitment especially childcare.

### Training gap

We also asked people what training they had already received and what they found useful. Almost a quarter of the respondents had received no training at all in the previous year. For those who had, one of the main benefits stated was networking with other research staff. While professional training courses such as good clinical practice and informed consent continue to be mandatory there is definitely an appetite for personal development.

Interestingly of the 44 people who cited leadership as the training area most important to their development and training needs, 59% were women. However, only 12 (27%) were clinical staff. There have been many studies on the importance of good medical leadership training. ^21^ There have been other studies that show that continuing medical education has the capacity to deliver high quality healthcare and to address many of the challenges in the health care environment. ^22^ Our study shows that there is an unfulfilled need for a range of training opportunities, particularly leadership training, for all translational researchers. The results of the survey will now help us to develop training as an integral part of career development pathways for our staff that meet their needs for professional and career advancement.

## CONCLUSIONS

Most researchers and research support staff in translational research settings like the NIHR BRCs want short training sessions, in a convenient location during the working day, preferably in the mornings. The training provided needs to be easily accessible, interactive and relevant. The most important areas for training include leadership skills, grant and fellowship writing and statistical applications. The biggest obstacle preventing translational researchers especially clinicians and nurses from doing training is work commitment.

## Data Availability

All data are reported in this article; hence, no additional data are available.

## Acknowledgements

The authors wish to thank all participants for taking time to complete the survey.

## Ethics statement

This study was an evaluation of the training and development service. We used the Health Research Authority (HRA) decision tool whether our study required NHS ethics approval. The HRA tool results suggested that our study would not be considered Research; hence, NHS ethics approval was not required and obtained. In addition, our retrospective application for ethics approval was reviewed by the Officer of the Oxford University Medical Sciences Interdivisional Research Ethics Committee, with reference to formally approved process and determined that the study would be classified as evaluation, rather than research, therefore does not require ethical review (CUREC Application: R77595/RE001, date August 26, 2021).

## Data availability statement

All data are reported in this article; hence, no additional data are available.

## Author’s contributions

KB devised and carried out the survey, did the initial analysis and wrote some parts of the manuscript. SGSS analysed the data, created visualisations and led drafting, revising and finalising of the manuscript. LRH reviewed and contributed to editing and redrafting the manuscript. VK reviewed the manuscript for critical intellectual input and acquired funds for open access publication of this paper.

## Appendix 1. NIHR Oxford BRC Training and Development Questionnaire

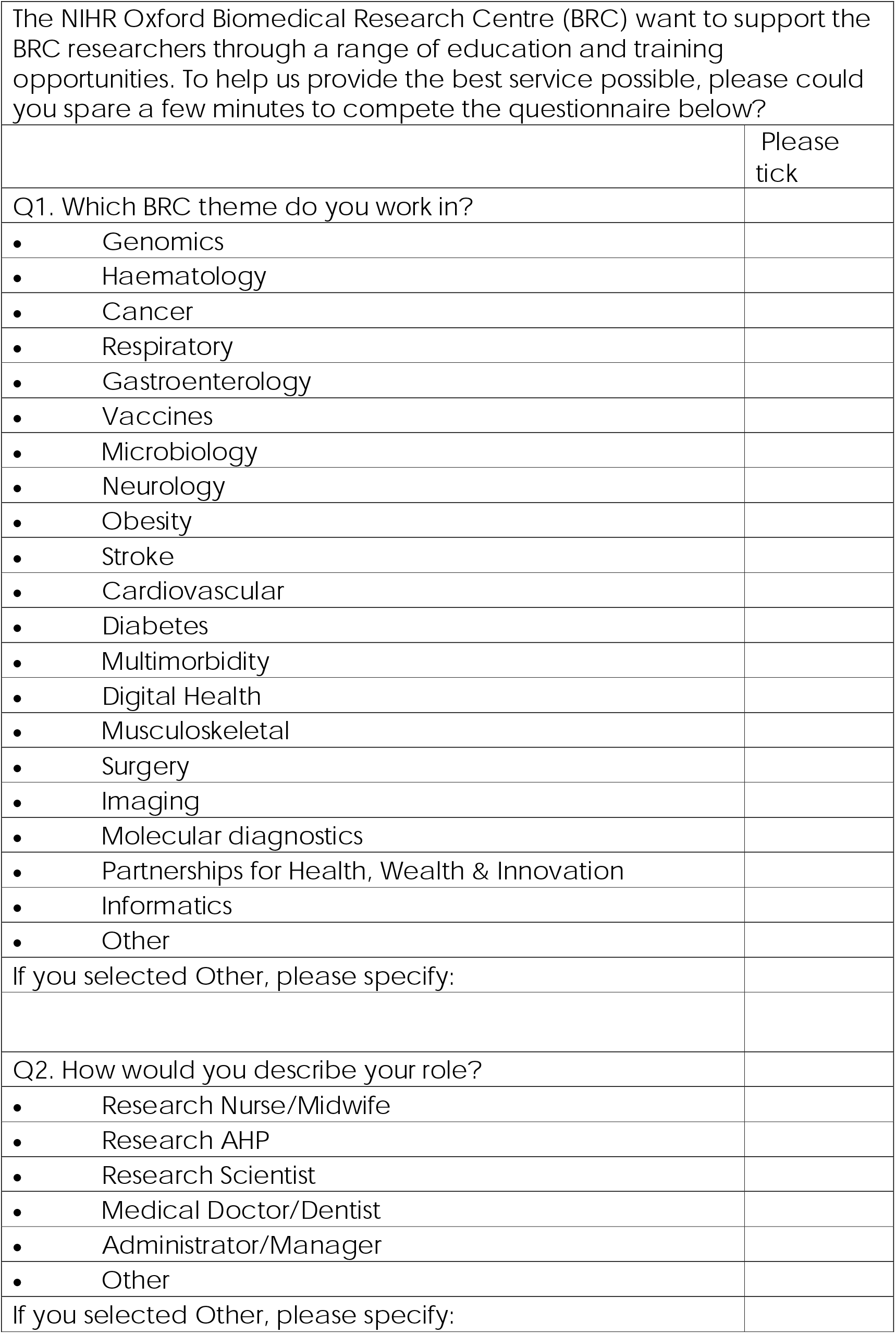
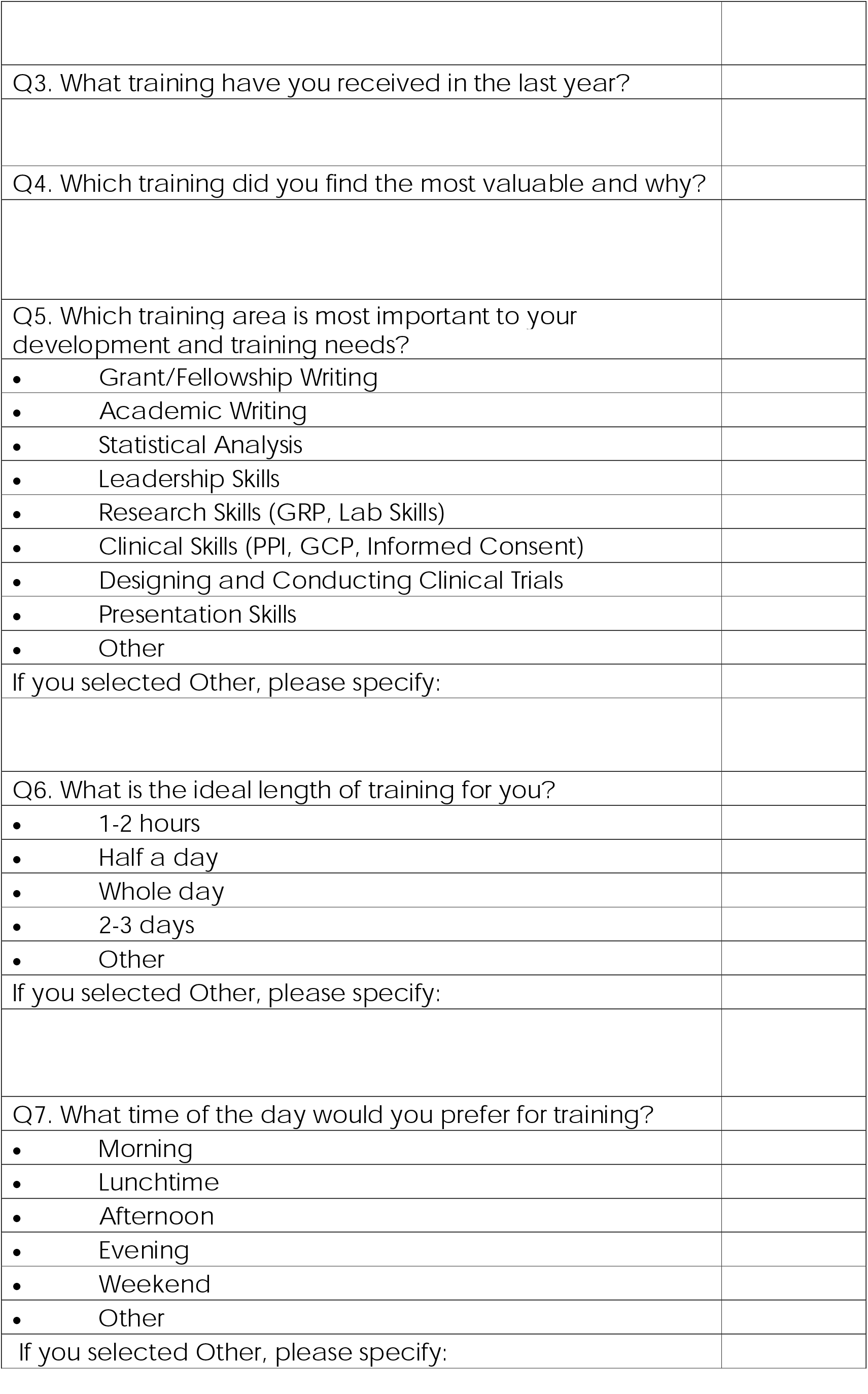
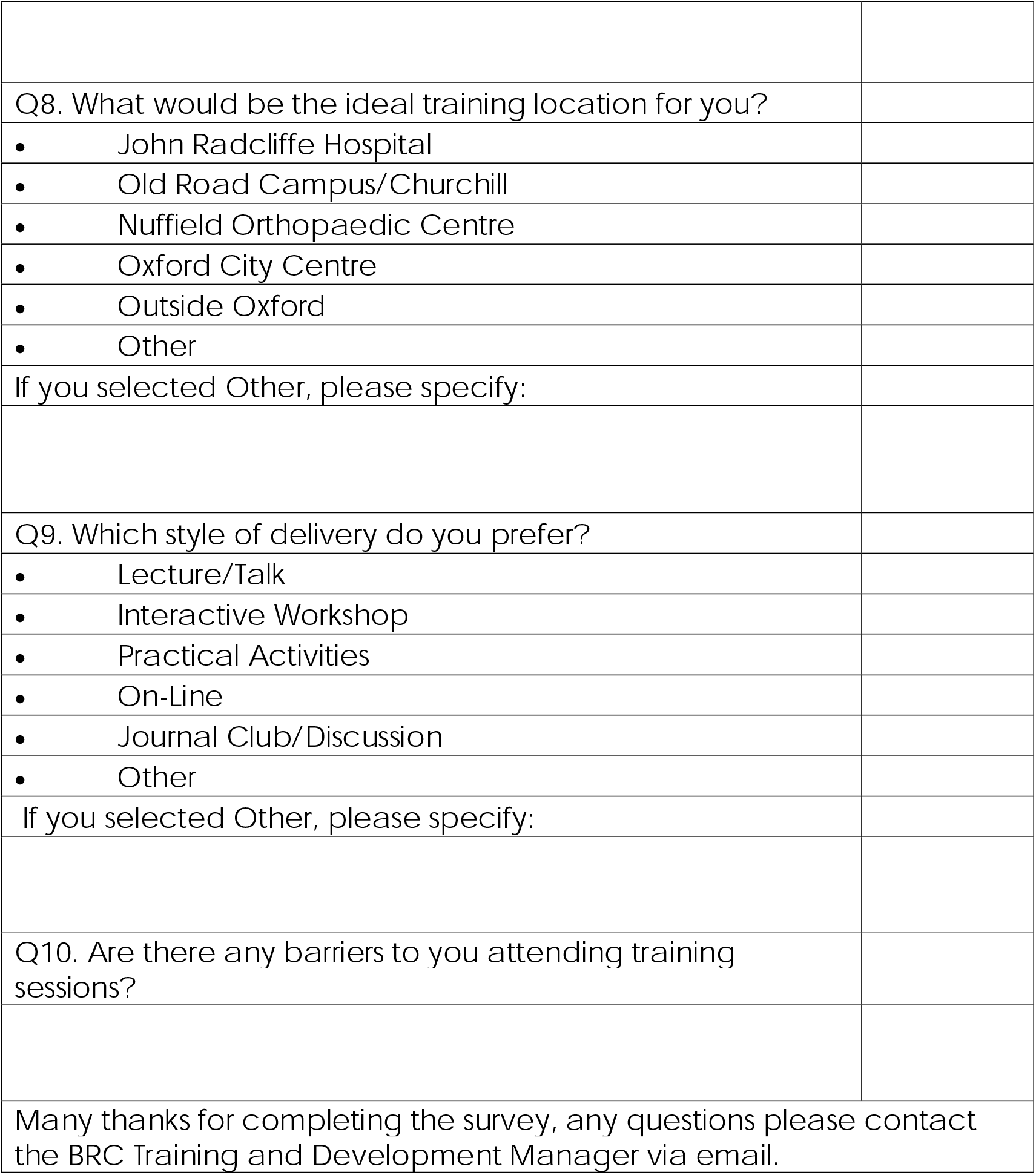

|  |  |
| --- | --- |
| The NIHR Oxford Biomedical Research Centre (BRC) want to support the BRC researchers through a range of education and training opportunities. To help us provide the best service possible, please could you spare a few minutes to complete the questionnaire below? |  |
|  | Please tick |
| Q1. Which BRC theme do you work in? |  |
| • Genomics |  |
| • Haematology |  |
| • Cancer |  |
| • Respiratory |  |
| • Gastroenterology |  |
| • Vaccines |  |
| • Microbiology |  |
| • Neurology |  |
| • Obesity |  |
| • Stroke |  |
| • Cardiovascular |  |
| • Diabetes |  |
| • Multimorbidity |  |
| • Digital Health |  |
| • Musculoskeletal |  |
| • Surgery |  |
| • Imaging |  |
| • Molecular diagnostics |  |
| • Partnerships for Health, Wealth & Innovation |  |
| • Informatics |  |
| • Other |  |
| If you selected Other, please specify: |  |
| Q2. How would you describe your role? |  |
| • Research Nurse/Midwife |  |
| • Research AHP |  |
| • Research Scientist |  |
| • Medical Doctor/Dentist |  |
| • Administrator/Manager |  |
| • Other |  |
| If you selected Other, please specify: |  |

|  |
| --- |
| Q3. What training have you received in the last year? |
| Q4. Which training did you find the most valuable and why? |
| Q5. Which training area is most important to your development and training needs? |
| • Grant/Fellowship Writing |
| • Academic Writing |
| • Statistical Analysis |
| • Leadership Skills |
| • Research Skills (GRP, Lab Skills) |
| • Clinical Skills (PPI, GCP, Informed Consent) |
| • Designing and Conducting Clinical Trials |
| • Presentation Skills |
| • Other |
| If you selected Other, please specify: |
| Q6. What is the ideal length of training for you? |
| • 1-2 hours |
| • Half a day |
| • Whole day |
| • 2-3 days |
| • Other |
| If you selected Other, please specify: |
| Q7. What time of the day would you prefer for training? |
| • Morning |
| • Lunchtime |
| • Afternoon |
| • Evening |
| • Weekend |
| • Other |
| If you selected Other, please specify: |

|  |
| --- |
| Q8. What would be the ideal training location for you? |
| • John Radcliffe Hospital |
| • Old Road Campus/Churchill |
| • Nuffield Orthopaedic Centre |
| • Oxford City Centre |
| • Outside Oxford |
| • Other |
| If you selected Other, please specify: |
| Q9. Which style of delivery do you prefer? |
| • Lecture/Talk |
| • Interactive Workshop |
| • Practical Activities |
| • On-Line |
| • Journal Club/Discussion |
| • Other |
| If you selected Other, please specify: |
| Q10. Are there any barriers to you attending training sessions? |
| Many thanks for completing the survey, any questions please contact the BRC Training and Development Manager via email. |

